# Toxigenecity and Virulence variations of *Pseudomonas aeruginosa* from out-patients hospitals in Southwest Nigeria

**DOI:** 10.1101/2025.01.07.25320112

**Authors:** Olusola Arinola Akinboboye, Olusola Abiodun Akingbade, Babatunde O Motayo, Eunice Folusho Olajumoke Akinleye, Aina Emmanuel Sesan, Idowu Adeyemi Samuel, Ede Dorathy Amuche, Nimotalahi Omotunde, Ede Dorathy Amuche

## Abstract

**Background:** Multidrug resistant (MDR) *Pseudomonas aeruginosa* isolates harboring genes for virulence and antibiotic resistance, have grown more prevalent lately. These strains pose a major threat to the general population, especially in tertiary care settings. There is a paucity of information on toxigenic and virulence diversity of multidrug resistant *P. aeruginosa* in Nigeria, hence, the need to characterize and determine the variations of the virulence genes.

**Methods:** Six hundred clinical samples from different anatomical sites were collected aseptically from Lagos University Teaching Hospital (LUTH), University of Medical Sciences, Ondo (UNIMED) and Federal Medical Centre, Abeokuta (FMC). *Pseudomonas aeruginosa* was isolated using cetrimide agar identified using biochemical tests. Antibiotic sensitivity was done by disc diffusion method. Protease, phospholipase C (lecithinase), caseinase and gelatinase presence were assayed for. Genomic DNA was extracted from *P. aeruginosa* isolates and screened for the presence of N-Acetylneuraminate synthase (NaN), Elastase B (*Las* B), Exotoxin A (ExoA), Exoenzyme S (ExoS) and Exoenzyme U (ExoU) virulence genes by PCR.

**Results:** Three hundred and sixty bacterial isolates identified from clinical samples are as follows: *Pseudomonas aeruginosa* (11.3%), *Escherichia coli* (18.0%), *Klebsiella pneumoniae* (14.3%), *Staphylococcus aureus* (10.2%), *Proteus mirabilis* (3.2%), *Streptococcus pnuemoniae* (2.3%*), Enterobacter aerogenes* (0.5%) and *Acinetobacter baumanni* (0.1%). Enzymes detected in the *P. aeruginosa* isolates were Phospholipase C (77.9%), caseinase (83.9%), gelatinase (98.5%) and protease (88.2%). The *P. aeruginosa* isolates were all resistant to ampicillin and cloxacillin; 26 (38.2 %) strains exhibited multidrug resistance. Virulence *Las*B elastase gene was detected in all 14 multi resistant *P. aeruginosa*, ExoA was detected in 5, ExoS in 4, ExoU in 5 and NaN in 4 isolates: Four (28.6%)

**Conclusion:** The study confirmed presence and variations of toxic genes in *Pseudomonas aeruginosa* isolated from all the three tertiary hospitals.

## INTRODUCTION

*Pseudomonas aeruginosa* has a number of virulence factors that contribute to its pathogenicity and antibiotic resistance. ExoU, a phospholipase, ExoY, an adenylate cyclase, and ExoS and ExoT, bifunctional proteins, are the four effector proteins that have been discovered [1]. The first type III secretion system toxins to be found, ExoT and ExoS, have 75% amino acid homology [2]. According to [3], ExoS is the main cytotoxin involved in colonization, invasion, and dissemination during infection. It specifically targets ras-like proteins, prevents eukaryotic cells from ingesting bacteria, slows DNA synthesis, and promotes death [2, 4]. ExoT specifically targets host kinases involved in phagocytosis and adhesion [4]. ExoY, an adenylate cyclase, breaks down the intracellular cAMP that induces certain cell types to round [5]. ExoU, the newest T3SS toxin discovered, has pronounced cytotoxic effects [4,5]. Exotoxin A (Exo A), which is produced by the Exo A gene and has the same mode of action as diphtheria toxin, inhibits protein synthesis through ADP-ribosylation of eukaryotic elongation factor 2. In the burn unit, intensive care unit, and among hospitalized patients, exotoxin A is a key virulence factor for *Pseudomonas aeruginosa* [6]. 90–95 percent of *Pseudomonas aeruginosa* have the gene that produces exotoxin A [7].

Another key enzyme that modulates bacteria pathogenesis in infected host is Exoenzyme S, which is encoded by the ExoS gene, induces apoptosis in macrophages and epithelial cells [8]. ExoS’s capacity to cause actin depolimerization and cytoskeleton disruption is correlated with its function in *Pseudomonas aeruginosa* pathogenesis [9]. In addition to blocking bacterial DNA synthesis and causing death, it is the main cytotoxin implicated in colonization, invasion, and dissemination during infection [10]. Exoenzymes U(ExoU)gene encodes a strong cytotoxin with phospholipase that may lyse host cell membranes [11]. According to 12Hotchkiss *et al*. (2018), the enzyme has been linked to septic shock and has been shown to worsen pneumonia symptoms and increase mortality, while ExoU expression has been linked to acute cytotoxicity [13]. Membranes of the infected host cells are disrupted by the presence of lipase activity [14]. Clinical infections result in the production of *Pseudomonas aeruginosa* elastase. Important biologic tissues and immune system components, such as immunoglobulin, serum complement factors, collagen, fibrin, and elastin, may be broken down by this enzyme. Las B is one of *Pseudomonas aeruginosa* ‘s most significant proteases, with a wide range of substrates including collagen, elastin, fibronectin, and lamins as well as immune and host defense molecules like gastric mucin, fibrin, transferrin, a-1 proteinase inhibitors IgG, and components of the complement pathway [15].

*Pseudomonas aeruginosa* type III produced toxins are significant virulence factors linked to clinically significant infections. These toxins are expressed by more than 80% of *Pseudomonas aeruginosa* strains that are isolated from acute infections, such as ventilator-associated pneumonia and sepsis [16]. Production of *Pseudomonas aeruginosa* toxins is associated with increased infection virulence and severity.

There is a paucity of data on the prevalence of *P aeruginosa* expressing these different virulent genes and invasive proteolytic enzymes among isolates recovered from patients in Nigeria. This gap in knowledge helped to motivate the design of this study. The aim of this study is to investigate the prevalence of MDR *P aeruginosa* isolates among patients with different types of infections in South West Nigeria, and determine the profiles of various virulent toxin and proteolytic enzyme expression.

## MATERIALS AND METHODS

### Study design and Study population

The research is a cross-sectional study of virulent properties of *P aeruginosa* isolates from South-Western, Nigeria. It was conducted at the following locations: Ondo State University of Medical Science Teaching Hospital, Laje Ondo, Ondo-State in South West Nigeria; Federal Medical Center, Abeokuta; and Lagos University Teaching Hospital, Mushin Idi-Araba, Lagos.

The Federal Medical Centre in Idi Aba, Abeokuta; Lagos University Teaching Hospital, Idi-Araba, Lagos; and Ondo State University of Medical Science Teaching Hospital, Ondo, South West, Nigeria, provided a total of 600 clinical samples which are urine, wounds, sputum, pus, ear swabs, and burns (two hundred each). A convenient sampling technique was adopted for sample recruitment. Ethical approval was obtained from the management of the three hospitals. Informed assent was obtained from patients, and clinicians who are involved in the management of the patients.

### Samples Collection

Using universal bottles and sterile swabs, the clinical samples (urine, wound swabs, sputum, ear swabs, pus, and burns) were aseptically taken from the patients. From every patient, two specimens were taken. The universal bottle and sterile swabs were appropriately labeled with the patients’ name, age, and sex.

### Culture and Identification of Isolates

The samples were inoculated into MacConkey agar, blood agar and cetrimide agar plates respectively and incubated aerobically at 37°C for 24 hours.

Numerous morphological and biochemical tests were performed on pure cultures of bacteria that had been isolated. Following that, Bergey’s Manual of Systematic Bacteriology was used to identify them. The experiments listed below were performed: Tests for Motility, Oxidase, Urease, Indole, Methyl Red, Vogue Proskauer, Citrate, Catalase, Coagulase, and Gram stain glucose, lactose, and sucrose fermentation.

### Antibiotics Susceptibility Testing

*Pseudomonas aeruginosa* isolates were tested for drug sensitivity and resistance using commercially available antimicrobial discs (Abtek Biological Ltd UK). The study used ten different antibiotics with varying disc concentrations: 25 µg for Amoxicillin (Amx), 25 µg for Cefuroxime (Cxm) Ceftazidime for 30 µg, 25 µg for Imipenem (Imp), 30 µg for Tetracycline (Tet), 10 µg for Streptomycin (Str), 5 µg for Cloxacillin (5 µg for Cxc), 25 µg for Gentamycin (Gen) and 15 µg for Erythromycin (Ery) and 30 µg for Ceftriaxone (Cef).

Every isolate underwent an antibiotic sensitivity test in accordance with the Kirby-Bauer disc diffusion method. The 0.5 Macfarland’s barium sulfate standard solution was used to compare the turbidity of the bacterial suspensions. Before applying the antimicrobial sensitivity discs, the standardized bacterial solution was injected onto Muller Hinton Agar (Lab M Laboratories, Mumbai, India) and allowed to dry for ten minutes. The test was conducted using 8mm-diameter antibiotic-impregnated discs. To ascertain the isolates’ antibiotic sensitivity, the width of the zone of inhibition was measured after incubation and compared with the zone diameter interpretation chart CLSI, 2020. As a control, *P. aeruginosa* ATCC 27853 standard strain was used.

### Protease production

Proteolytic activity was assessed using the methodology described by [17]. 15 mg of hide powder were treated with 3 ml of supernatant in 10 mM Tris HCl buffer (pH 7.5) at 37°C for 1 hour while being vigorously shaken. Centrifugation at 3000 g for 10 min was used to remove the undissolved substrate, and the supernatants’ protease activity was measured at 59

### Detection of phospholipase C (lecithinase)

*Pseudomonas aeruginosa* isolates were seeded on the egg yolk agar medium’s surface and incubated at 37 °C for 24 to 48 hours, according to [18]. The emergence of colonies surrounded by an opacity zone indicates a positive outcome.

### Detection of caseinase

*Pseudomonas aeruginosa* isolates were reportedly injected on the surface of milk agar medium and incubated at 37 °C for 24 hours, according to [19]. A zone of clearing underneath and around the growth, resulting from clear and cut responses that appear within 24 to 48 hours, would suggest a positive outcome.

### Detection of gelatinase

*Pseudomonas aeruginosa* culture was incubated at 37 °C for seven days. The tube was periodically removed from the incubator and held at 4 °C for half an hour before the results were read. As long as the gelatin was still liquid, a positive outcome was seen.

### Chromosomal DNA Extraction

Following growth on TSA medium, fourteen multidrug-resistant plasmid-mediated *Pseudomonas aeruginosa* isolates were cultured overnight. Purified genomic bacterial DNA was extracted using a genomic DNA micro kit (QIAGEN, QIAamp® - USA) in accordance with the manufacturer’s instructions. This acts as the DNA template. Using a NanoDrop 2000 spectrophotometer, the concentration of the eluted DNA was determined.

### Polymerase Chain Reaction

The polymerase chain reaction (PCR) was used to target the genes N-Acetylneuraminate synthase (NaN 1), Elastase B (Las B), Exotoxin A (exoA), and Exoenzyme S and U (exoS and exoU) in the pure DNA of each distinct *Pseudomonas aeruginosa*. 25 ng of DNA template, 10 mM Tris-HCl, 50 nmol KCl, 1.5 mM MgCl2, 200 µM dNTP (Fermentas), 12.5 pmol of each primer, 1 U Taq DNA polymerase (Fermentas), and 5 µL PCR buffer 10X were used in the 50 µL PCR experiment. One cycle of 94°C for three minutes started the reaction, which was then followed by thirty cycles at 94°C for thirty seconds, 55°C for one minute, 72°C for 1.5 minutes, and a final elongation step at 72°C for five minutes.

**Table 1.**
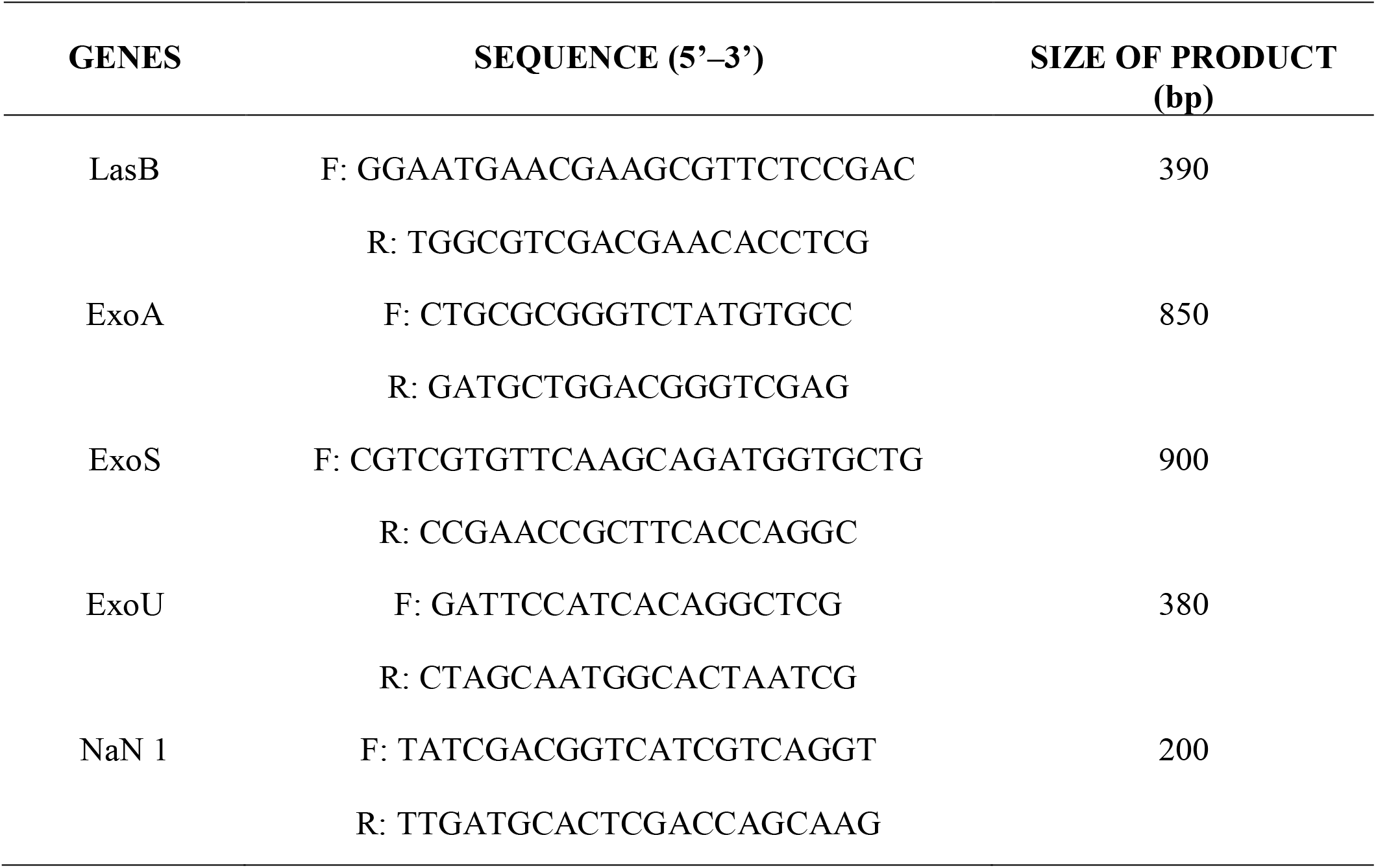
Oligonucleotide primers of virulence genes.

### Statistical Analysis

For analysis, the data were moved to a Microsoft Excel spreadsheet (Microsoft Corp., Redmond, WA). Data analysis was carried out using R Studio (https://www.r-project.org) to identify distribution and association between the virulence genes, antimicrobial resistance, and occurrences of bacteria according to age and gender *Pseudomonas aeruginosa* isolated from clinical samples.

## RESULTS

In this investigation, 600 clinical samples were obtained; they included sputum (15.4%), urine (46.5%), burns (5.0 %), ear swabs (7.3%), pus (8.3%), and wound swabs (17.5 %), a visual distribution of the samples collected is shown as a donut plot in Figure 1A. Out of the six hundred clinical samples cultured, Three hundred and sixty samples yielded bacterial growth while two hundred and forty samples had no growth.

**FIGURE 1A.**
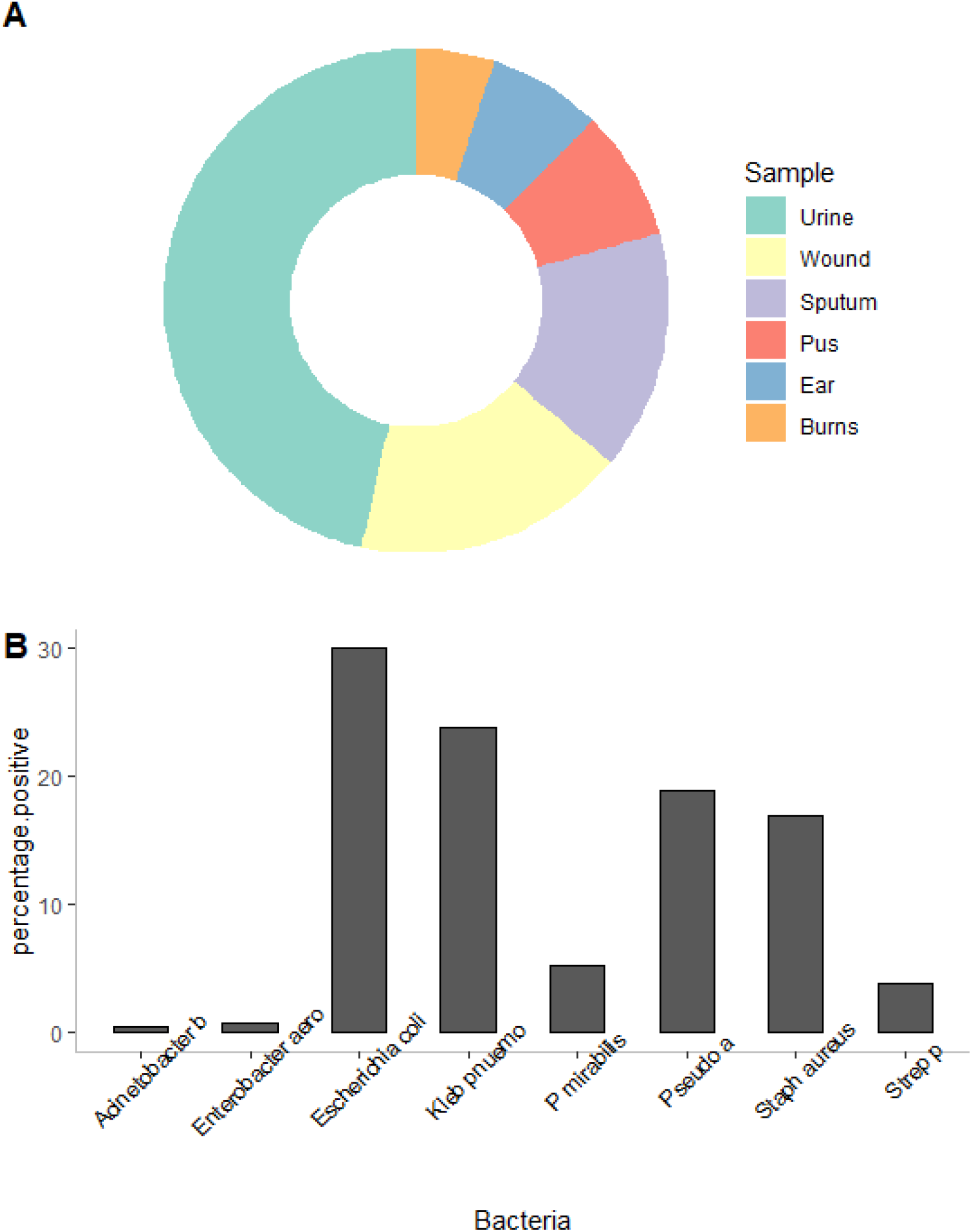
Donut plot showing the distribution of sample type utilized for *Pseudomonas aeruginosa* isolation in this study. 1B. Bar chart showing the percentage distribution of different bacteria species isolated during the study.

Figure 1B shows the distribution of bacteria species isolated in this study, *Staphylococcus aureus* (61), *Proteus mirabilis* (19), *Streptococcus pnuemoniae* (14), *Enterobacter aerogenes* (3), and *Acinetobacter baumannii* 10.2%, 3.2%, 2.3%, 0.5, and 0.2%.

There was no statistically significant difference in the proportion of *Pseudomonas aeruginosa* isolated from clinical samples taken from female 29 patients (4.0%), compared to male 39 patients (6.5%). Table 2 shows that patients between the ages of 31 and 40 had the greatest incidence of *Pseudomonas aeruginosa* isolates 19 (3.2%) from their samples, while patients under the age of 10 and those over 50 had the lowest (1.2%).

**Table 2.** Distribution of *Pseudomonas aeruginosa* isolated from clinical samples in relation to sex and age.

| Sex | Total examined (%) | Number (%) yielded growth of *Pseudomonas aeruginosa* |
| --- | --- | --- |
| Male | 255(42.5) | 29(4.0) |
| Female | 345(57.5) | 39(6.5) |
|  | 600(100) | 68(11.3) |
| Age group |  |  |
| ≤10 | 25(4.2) | 7(1.2) |
| 11 – 20 | 92(15.3) | 9(1.5) |
| 21 – 30 | 113(18.8) | 12(2.0) |
| 31 – 40 | 173(28.8) | 19(3.2) |
| 41 -50 | 95(15.8) | 14(2.3) |
| ≥ 51 | 102(17.0) | 7(1.2) |
| **Total** | **600(100)** | **68(11.3)** |

p > 0.05

Figure 2A represents the geographical distribution of *pseudomonas aeruginosa* isolated from the different States of Nigeria where the study was conducted, from our data, Ondo State recorded the highest distribution of *P. aeruginosa*, followed by Ogun State and lastly Lagos State. Figure 2B shows a facetted pie chart distribution of percentage of isolates producing proteolytic enzymes by state, with each pie char representing each type of enzyme. The sectors in each chart represents the percentage distribution of isolates by State of isolation. From the charts Ogun State recorded the highest level of enzyme production in all 3 enzyme types, with Lagos State recorded the least enzyme production among the isolates (Figure 2B).

**FIGURE 2A.**
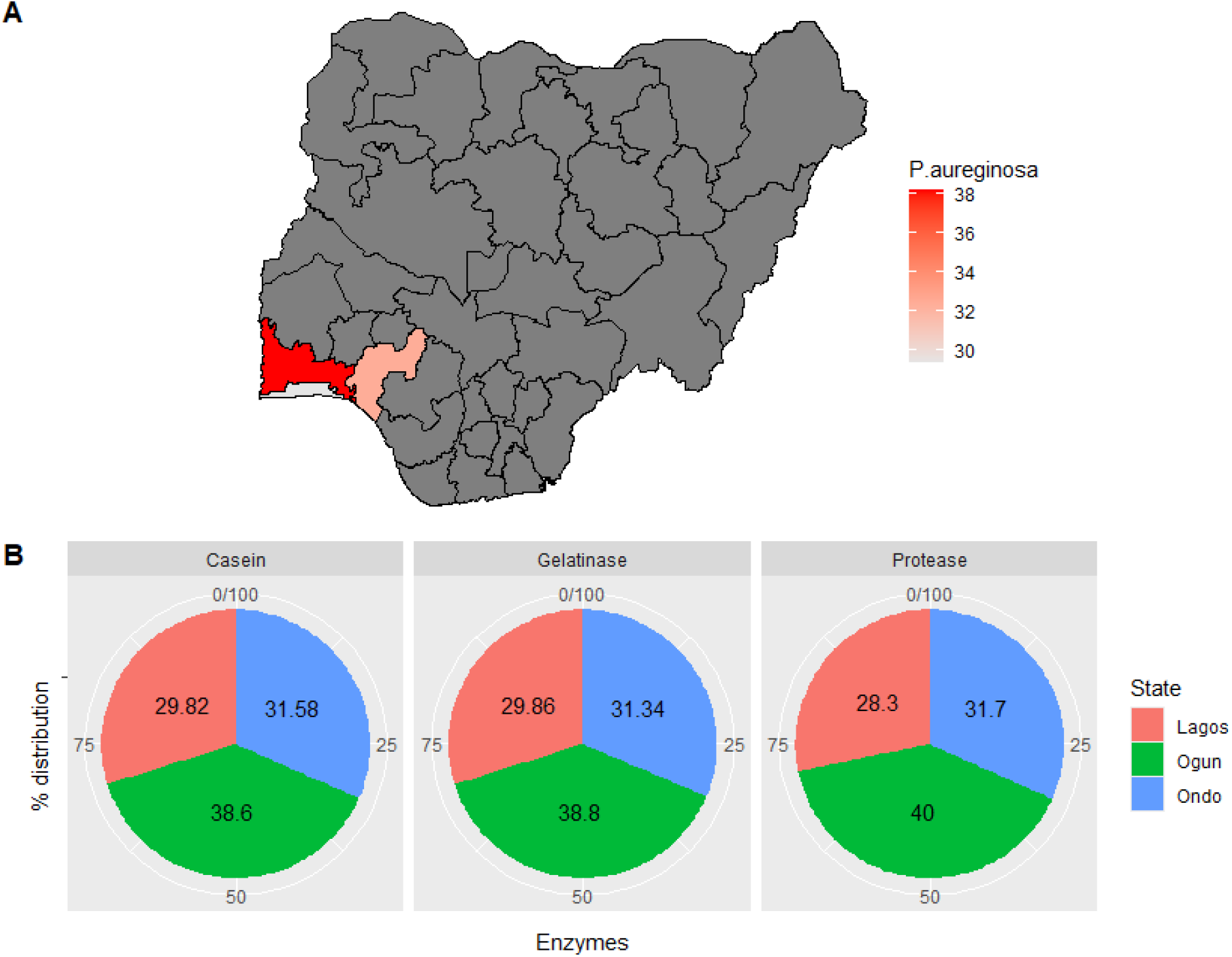
Geographic distribution of *Pseudomonas aeruginosa* among the 3 states of Oyo, Ogun and Lagos, Southwest Nigeria where the study sites were located. 2B, Pie charts showing State-wise distribution of proteolytic enzyme producing *Pseudomonas aeruginosa* isolates.

The antibiotics susceptibility showed that the 68 different *Pseudomonas aeruginosa* isolates tested, were resistant to one or more antibiotics used. Sixty-eight (100%) of the isolates tested, were resistant to ampicillin and cloxacillin respectively, 67(98.5%) were resistant to erythromycin, while fifty-nine (86.8%) were resistant tetraycline. Fifty-two (76.5%) of the isolates were resistant to ofloxacin, 48 (70.6%) to gentamycin while 31 (45.6%) were resistant to ceftriaxone and cefuroxime respectively. Twenty -one (30.9%), 18(26.5%) and 10 (14.7%) were resistant to cefepime. Ceftazidime and imipenem respectively Figure 3.

**FIGURE 3.**
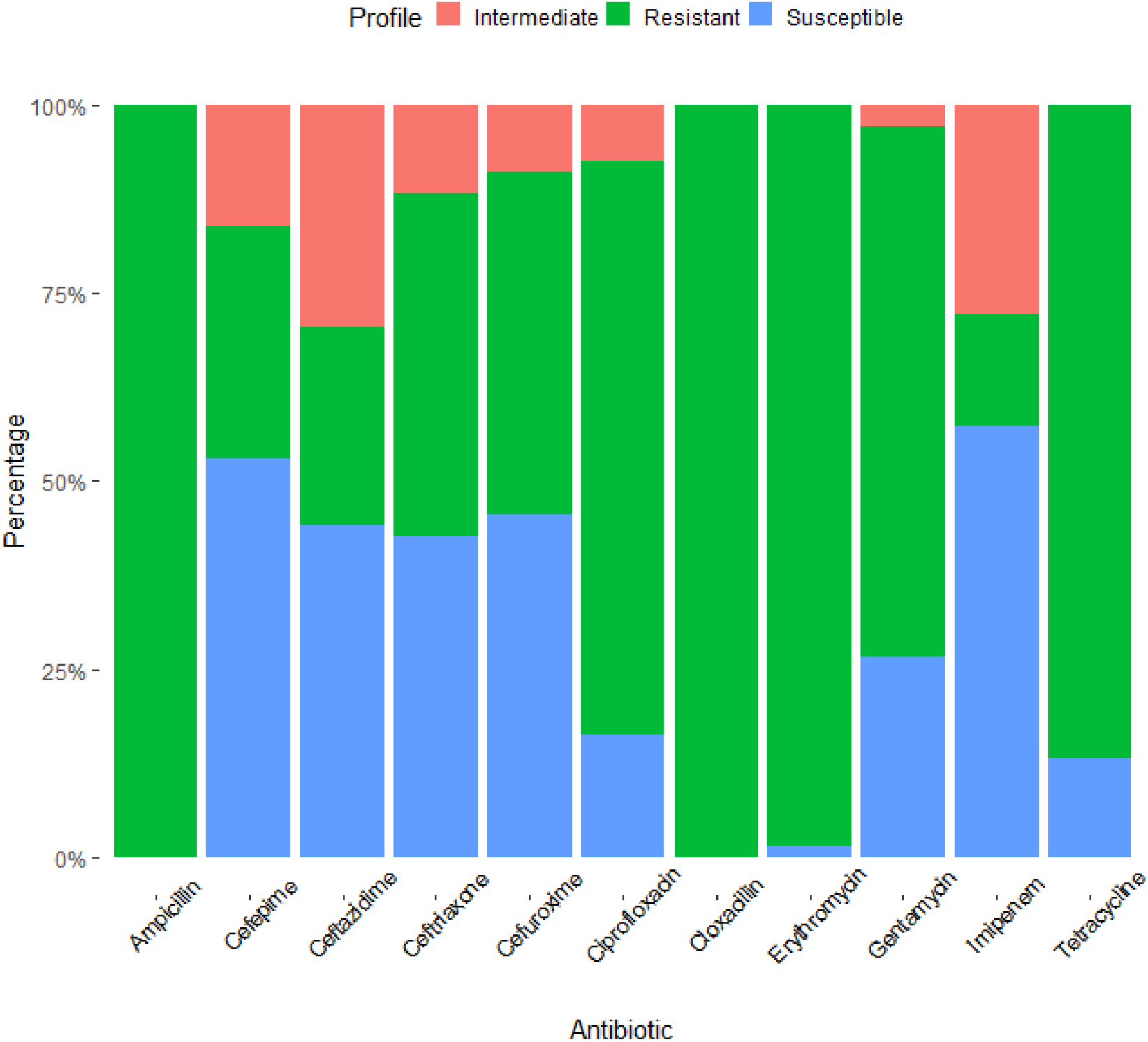
Stacked bar chart showing percentage distribution of antibiotic susceptibility profiles of Pseudomonas aeruginosa isolates tested against various types of antibiotics, the legend shows the color code for the susceptibility profile.

All fourteen of the multidrug-resistant Pseudomonas aeruginosa isolates on plates 1a and 1b had the LasB elastase virulence genes. Five of the multidrug resistant Pseudomonas aeruginosa isolates (Figure 4A plates I and II) had the virulence Exo A gene, whereas four of the multidrug resistant Pseudomonas aeruginosa isolates had the virulence Exo S gene. Four of the multidrug-resistant Pseudomonas aeruginosa isolates had virulence NaN 1, while five of the multidrug-resistant isolates on plate 5 had virulence Exo U, Figure 4A plate V.

**FIGURE 4A.**
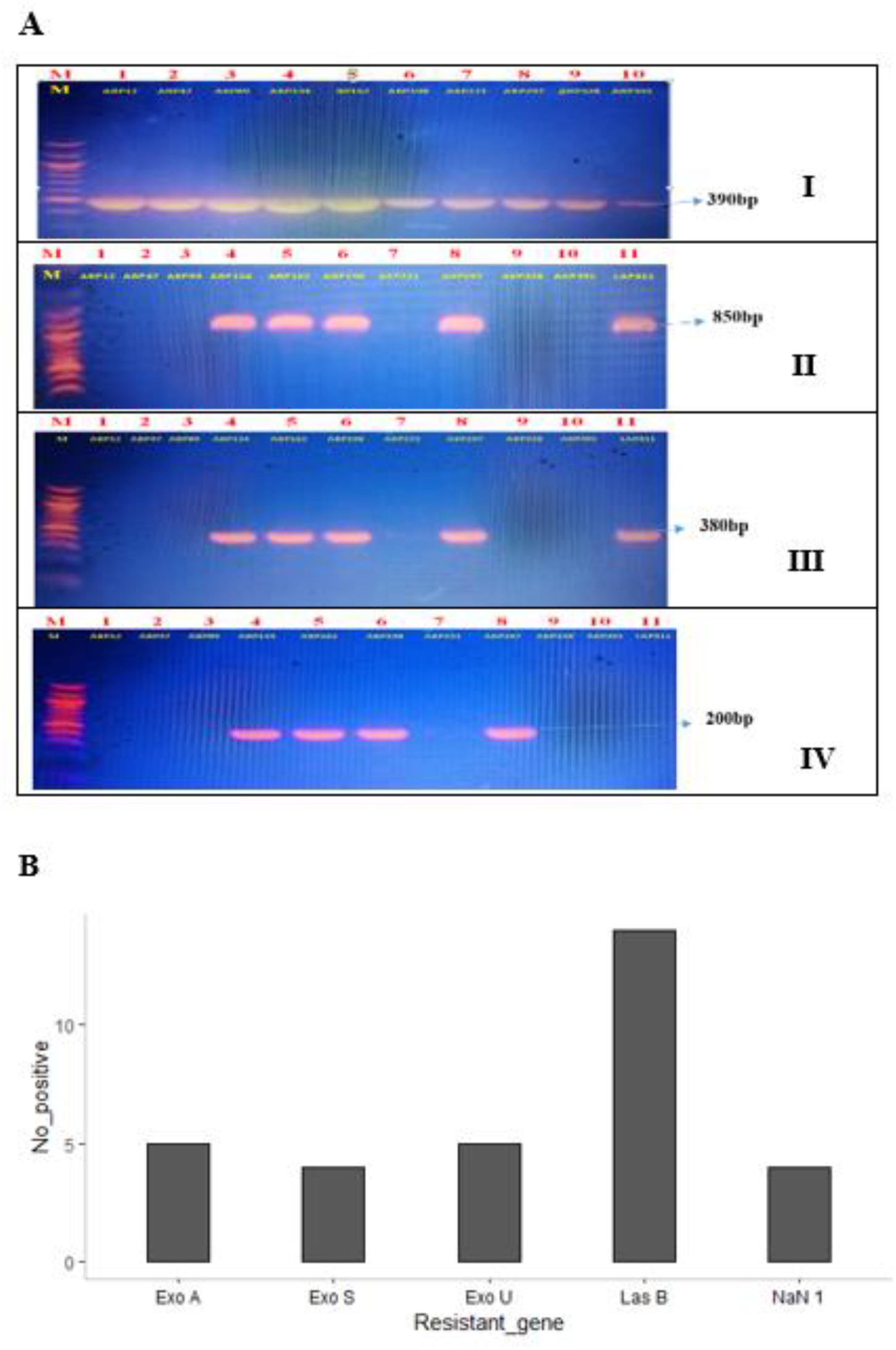
Gel electrophoresis panels showing positive PCR reactions for Exo A gene I and II, Exo S gene III and NaN I (IV), the DNA ladder is seen on the first row as banded lines. FIGURE 4B. Bar chart showing the positivity rate of the various virulence genes screened among drug resistant *Pseudomonas aeruginosa* from South West, Nigeria.

## DISCUSSION

The adaptable bacteria *Pseudomonas aeruginosa* produces a variety of severe opportunistic infections in people with significant underlying medical problems. Six hundred clinical samples were used in this investigation to identify *Pseudomonas aeruginosa* isolates. *Pseudomonas aeruginosa* was present in 11.3% of cases. Since *Pseudomonas aeruginosa* may express a wide range of virulence factors, adapt to changes in its environment, and develop antibiotic resistance, it is well recognized that infections with this pathogen are linked to high rates of morbidity and death. Nowadays, one of the most common nosocomial pathogens is *Pseudomonas aeruginosa*, and because of antibiotic resistance, infections caused by this bacterium are often challenging to treat. Only 38.2% of all *Pseudomonas aeruginosa* isolates were found to be multidrug resistant, according to the research. Comparing this to the 63.2% multidrug resistant *Pseudomonas aeruginosa* published by [20], it is less. The antibiotic classes that were selected to treat the isolated *Pseudomonas aeruginosa* strain were imipenem, tetracycline, aminoglycosides, macrolides, cephalosporins, quinolones, and penicillin. The efficiency of several antibiotics against *Pseudomonas aeruginosa* has been investigated in a variety of research [21-23]. Every single *Pseudomonas aeruginosa* that was found was completely resistant to ampicillin and cloxacillin, two of the most widely used medicines in the penicillin family. 90% and 88.2% of South West Nigerians, respectively, were found to be cloxacillin and ampicillin resistant [24]. Among the first class of antibiotics used to treat various illnesses are ampicillin and cloxacillin. Concerningly, *Pseudomonas aeruginosa* has a strong resistance to them. All isolates from the three zones under study shown strong resistance to ciprofloxacin, tetracycline, and erythromycin in *Pseudomonas aeruginosa*. The frequency of multidrug resistant *Pseudomonas aeruginosa* often varies significantly across communities and hospitals in the same location, as well as among numerous patients and populations in the same hospital [22,25,26]. In this investigation, the β-lactam antibiotics cefepime and ceftazidime, shown an excellent sensitivity pattern against *Pseudomonas aeruginosa*. The sensitivity of cefepime, a fourth-generation cephalosporin, to *Pseudomonas aeruginosa* was 69.1%. This is compared to 56% resistant reported in comparable research from Iran [27]. In this trial, the sensitivity of the third-generation cephalosporin ceftazidime was 73.5%. According to [28], drugs have high sensitivity against *Pseudomonas aeruginosa*. The outcome also agrees with previous research [29], although it disagrees with another similar study [30] that found that *Pseudomonas aeruginosa* was 96.6% resistant to ceftazidime. The study’s findings regarding the high sensitivity of cephalosporins run counter to the assertion made by previous workers [31] that *Pseudomonas aeruginosa* is naturally resistant to all β-lactam antibiotics, including broad spectrum cephalosporins, primarily due to the incredibly low permeability of their cell wall. Cefepime and ceftazidime showed a high sensitivity against *Pseudomonas aeruginosa* in this investigation; however, this might be due to other variables that allow the antibiotics to pass through the isolates’ poor permeability. Several regulatory and virulence factors of *Pseudomonas aeruginosa* contribute in the process of bacterial infection. The virulence genes that *Pseudomonas aeruginosa* harbors, including Las B elastase, Exotoxin A (Exo A), Exoenzymes S (ExoS), Exoenzymes U (ExoU), and N-Acetylneuraminate synthase (NaN 1), were identified using the polymerase chain reaction method. In all fourteen (100%) of the plasmid-mediated, multidrug-resistant *Pseudomonas aeruginosa* samples examined, LasB genes were found. The most prevalent pathogenicity gene (100%) among the isolates evaluated was this LasB gene [32, 33], which encoded for elastase. In another study conducted in South Africa, the LasB gene was found in 10.9% of the *Pseudomonas aeruginosa* that was investigated. [33] claim that the Las system controls the synthesis of a number of virulence agents, including alkaline protease, exotoxin A, and elastase. Furthermore, the Las system positively controls the Rhl system. A few virulence factors, including alkaline protease, biofilm formation that promotes the manufacture of pel polysaccharide, elastase, rhamnolipid, pyocyanin, and HCN, are induced by the rhl system [34]. 35.7 percent of the 14 multidrug-resistant *Pseudomonas aeruginosa* samples that were screened tested positive for Exo A. This is less than the 96.6 and 90.7% of samples that were reported by [10, 35], respectively. One possible explanation for the 64.3% of *Pseudomonas aeruginosa* that tested negative for Exo A might be because certain strains of the bacteria naturally lack this gene. When isolates examined had the Nan1 genotype, the frequency of the Nan1 gene was much lower (28.6%). 35.7% of the multidrug-resistant *Pseudomonas aeruginosa* isolates have ExoU genes, which is less than the 97% reported by [36]. The gene encoding for exoenzyme (ExoS) was discovered to be present in a larger proportion of *Pseudomonas aeruginosa* isolates (28.6%) according to research conducted in Iran [1]. It’s possible that some of the isolates that tested negative for one or more virulence genes had their genes mutated away. ExoS is present in invasive strains, but ExoU seems to be present in cytotoxic strains [37]. ExoU is more than 100 times more cytotoxic than ExoS [38]. Resistance develops when strains with virulence factors—such as toxin genes—cause more illness symptoms and, therefore, are exposed to more antimicrobial drugs. The five virulence genes, lasB/exo A/exoS/exoU/NAN1, were only present in 28.5% of the isolates, whereas 7.1% of the isolates carried three of the genes.

## CONCLUSION

The results confirmed that *Pseudomonas aeruginosa* was isolated from clinical samples and comprised the virulence genes NaN 1, LasB, ExoA, Exo S, and Exo U. Most of the *Pseudomonas aeruginosa* isolates in all three locations were found to be resistant to ampicillin, amoxicillin, cloxacillin, and other commonly used antibiotics.

## Data Availability

All data produced in the present work are contained in the manuscript

